# Sociodemographic Characteristics of Missing Data in Digital Phenotyping

**DOI:** 10.1101/2020.12.29.20249002

**Authors:** Mathew V Kiang, Jarvis T Chen, Nancy Krieger, Caroline O Buckee, Monica J Alexander, Justin T Baker, Randy L Buckner, Garth Coombs, Janet W Rich-Edwards, Kenzie W Carlson, Jukka-Pekka Onnela

## Abstract

The ubiquity of smartphones, with their increasingly sophisticated array of sensors, presents an unprecedented opportunity for researchers to collect diverse, temporally-dense data about human behavior while minimizing participant burden. Researchers increasingly make use of smartphone applications for “digital phenotyping,” the collection of phone sensor and log data to study the lived experiences of subjects in their natural environments. While digital phenotyping has shown promise in fields such as psychiatry and neuroscience, there are fundamental gaps in our knowledge about data collection and non-collection (i.e., missing data) in smartphone-based digital phenotyping. Here, we show that digital phenotyping presents a viable method of data collection, over long time periods, across diverse study participants with a range of sociodemographic characteristics. We examined accelerometer and GPS sensor data of 211 participants, amounting to 29,500 person-days of observation, using Bayesian hierarchical negative binomial regression. We found that iOS users had higher rates of accelerometer non-collection but lower GPS non-collection than Android users. For GPS data, rates of non-collection did not differ by race/ethnicity, education, age, or gender. For accelerometer data, Black participants had higher rates of non-collection while Asian participants had slightly lower non-collection. For both sensors, non-collection increased by 0.5% to 0.9% per week. These results demonstrate the feasibility of using smartphone-based digital phenotyping across diverse populations, for extended periods of time, and within diverse cohorts. As smartphones become increasingly embedded in everyday life, the insights of this study will help guide the design, planning, and analysis of digital phenotyping studies.

## Introduction

The ubiquity of personal digital devices has resulted in a unique opportunity to collect and analyze unprecedented amounts of data, providing researchers with a promise of a more nuanced understanding of human behavior than ever before. This trend continues to accelerate as internet-connected personal devices become more prevalent, accessible, and embedded in everyday life. According to a recent study, over half of the world population has internet access.^1^ Cellular phones outnumbered humans globally in 2014,^2^ and current projections estimate over six billion smartphones to be in circulation by the end of 2020,^3^ making smartphones the fastest growing technology in history.^4^ In the United States, smartphone ownership is currently estimated at 77%, up from just 35% in 2011.^5^

Leveraging the resulting data deluge to understand human behavior in a more granular and precise manner, public health researchers have created the field of “digital epidemiology.” Defined as health-related research using data generated outside of the health system and for non-health-related research purposes,^6,7^ digital epidemiology has advanced our understanding of how health and collective human behavior interact. For example, mobile phone data from telecommunications providers have been used to quantify the impact of human mobility on malaria transmission,^8^ emerging dengue epidemics,^9^ and access to health care.^10^ Google search queries, combined with historical case data, have demonstrated predictive power for tracking epidemics of both influenza^11^ and dengue.^12^ Similarly, social media data have been used to predict Zika incidence^13^ and city-level influenza emergency department visits.^14^

While digital epidemiology focuses on patterns of collective human behavior, “digital phenotyping” seeks to learn about individual-level human behavior. We have previously defined digital phenotyping as “the moment-by-moment quantification of the individual-level human phenotype *in situ* using data from personal digital devices,” in particular smartphones.^15–17^ As with any scientific inquiry, measurement is vital, and these personal digital devices provide an unprecedented opportunity for precise measurement of human behavior, at fine spatiotemporal resolution, using existing consumer grade devices across large, diverse samples. This pairing of individual-level data collection and analysis creates a nuanced view of the participant’s “digital phenotype,”^18^ allowing researchers to understand the lived experience of subjects. The goal of digital phenotyping is to provide more precise social, behavioral, and cognitive phenotypes for developing a better understanding of various diseases, potentially leading to the establishment of new disease subtypes in fields such as psychiatry and neurology. These more precise phenotypes could enable early and accurate detection of diseases, thus advancing the goals of precision medicine, and monitor treatment response in an unobtrusive manner while facilitating measurement-based care at scale.

While still nascent, digital phenotyping has shown significant promise, especially in the field of mental health.^19–21^ For example, several studies have found a link between individual-level mobility, estimated from smartphone GPS sensor data, and depressive symptoms.^22^ Among schizophrenia patients, digital phenotyping has been shown to be acceptable to patients and potentially feasible for use in clinical practice,^23^ predictive of schizophrenic relapse in a small pilot study,^24^ and capable of providing scalable and affordable sleep monitoring.^25^ Additionally, digital phenotyping has begun to branch out to other areas of population health research: understanding the daily behaviors of healthy undergraduate students,^26^ evaluating the risk of disordered eating among women with and without histories of childhood trauma and food insecurity, monitoring patient recovery after cancer surgery,^27^ and providing enhanced medical care within a cohort of patients with advanced cancer.^28^ However, researchers have also called for a better understanding of how these data are collected,^29^ greater emphasis on methodology and techniques for analyses of these data rather than just on the collection itself,^16,30^ and, as with any new area of research, establishing more ethical standards and guidelines for data collection.^31^

While many platforms exist for collecting data from smartphones, we focus on studies using Beiwe, a research platform for smartphone-based digital phenotyping. The development of Beiwe started in 2013, and the first version of the platform was introduced in 2016 and is described in detail elsewhere.^15^ Briefly, Beiwe is a scalable, globally deployable, cloud-based data collection and data analysis platform designed for smartphone-based digital phenotyping in biomedical settings. Some of its distinguishing features are the ability to collect raw sensor data rather than pre-packaged data summaries, support for Android and iOS devices, emphasis on reproducibility of research through sharing of study configuration files, and full back-end integration with the Forest data analysis library that consists of statistical and machine learning methods specifically developed for analyzing smartphone data. Beiwe has been released under the 3-clause BSD open source license, which enables researchers to modify and expand the capabilities of the platform to meet their own scientific needs (Supplementary Information Text S1). Among other features, the platform allows investigators to specify which data streams are collected, how frequently they are sampled, and how frequently the data are uploaded to the server. Data are encrypted while buffered on the phone awaiting upload, during transit, and while at rest on the server. The support for both iOS and Android devices covers an estimated 99% of the U.S. smartphone market.^32^

Despite the potential for scalable, affordable, intensive data collection with a beneficial impact on medicine and public health, many fundamental questions about digital phenotyping data collection remain unanswered at this early stage of the field. For example, previous research has noted differences in smartphone mean duration of usage by gender and primary purpose of phone usage by age.^33^ Other researchers have documented differences in the sequencing of smartphone application usage^34^ and perceptions of battery life.^35^ While the demographic differences in phone usage are clear, albeit under-researched, it remains unclear how these demographic differences may affect levels of missingness in smartphone-based digital phenotyping data collection. This is an important unresolved question in the field because missingness in digital phenotyping data can undermine the usefulness of many medical or public health applications.

Missing data in digital phenotyping can divided into two categories: (1) *missingness by design* and (2) *missingness due to sensor non-collection*. Missingness by design is an intended result of the sensor sampling schedule as configured by the investigator. For example, to preserve phone battery, at the design stage an investigator might configure the GPS sensor to collect data for 1 minute every 10 minutes. In contrast, missingness due to sensor non-collection results from technological and human factors. For example, a participant may forget to charge their phone, disable the GPS, or uninstall the study application. The phone’s operating system may also limit sensor access during specific scenarios due to performance considerations. Because the technological factors causing sensor non-collection are usually proprietary and therefore unknown to the investigator, identifying sensor non-collection and characterizing its extent is crucial so that the investigator, at a minimum, can quantify the resulting additional uncertainty in downstream data analyses, and can also consider imputing missing data. For smartphone applications that alternate sensor sampling between an on-cycle (sensor collets data) and off-cycle (sensor does not collect data), the expected data volume is known at the design stage, which enables one to easily diagnose sensor non-collection. In the above example, collecting data from the GPS sensor every 10 minutes for 1 minute at a time leads to a regular 10% sampling coverage of any time period, resulting in 2.4 hours of data for every 24-hour period, for example. While outside the scope of this paper, we note that missingness due to sensor non-collection can be further divided into subtypes, such as missing completely at random, missing at random, and not missing at random, and distinguishing between these missing data mechanisms is important at the data analysis stage.^36^

This study focuses on sensor non-collection and seeks to address four fundamental questions about this type of missingness in digital phenotyping data collection from accelerometer and GPS sensors: (1) What is the expected rate of sensor non-collection for accelerometer and GPS in digital phenotyping studies? (2) To what extent does the rate of sensor non-collection vary over the study period? (3) How are rates of sensor non-collection correlated with phone type or common demographic characteristics of participants, such as gender, education, or age? (4) What is the individual-level variability of sensor non-collection? As far as we know, this is the first systematic investigation of these issues in a cross-diagnostic cohort in digital phenotyping.

## Results

We analyzed the timestamps of accelerometer and GPS measurements collected in six different studies, conducted in 2015–2018, with a combined total of 211 participants (Figures 1, 2, and S1) using the Beiwe Research Platform (Table 1). In all, there were over 8.3 billion measurements (8.1 billion individual accelerometer measurements and 113 million GPS individual measurements) collected in over 81 million measurement groupings over the course of more than 29,500 person-days of observation (Table S1). For all analyses reported in this paper, we used only timestamps of each measurement and not the measurement itself. Identifying information, such as GPS coordinates, were not necessary for the objectives of this study and thus all sensor measurements were removed before analysis. In addition to timestamps, we collected self-reported demographic information about participants in most of these studies (Table 1). These self-reported demographic data include gender, age, educational attainment (highest completed degree), and race/ethnicity (non-Hispanic White, non-Hispanic Black, Asian, American Indian/Alaska Native, other/Hispanic). Overall, among the 211 participants, the average age at the beginning of each study was 25.4 years (SD 10.8), most were men (66%), most had at most a high school education (67%), and 55% were non-Hispanic White, with the next two most common racial/ethnic groups being Asian (17%) and Black (14%).

**Table 1.** Study demographic characteristics. General sociodemographic characteristics of each study and across all studies. Studies A, C, E, and F consisted of healthy undergraduate students from Harvard College. Study B consisted of patients known to be at risk for mania and psychosis from McLean Hospital in Massachusetts. Study D consisted of healthy female nurses from the Nurses’ Health Study 3. In parentheses, the Total column shows the row percent relative to the entire sample except for the age row where it shows the sample standard deviation of age in years.

|  | Study A | Study B | Study C | Study D | Study E | Study F | Total (%) |
| --- | --- | --- | --- | --- | --- | --- | --- |
| Participants, N | 16 | 11 | 12 | 59 | 39 | 74 | 211 (100%) |
| Mean (SD) age, y | 19.4 (1.2) | 31.5 (9.5) | 20.4 (1.5) | 41.1 (6.3) | 18.4 (0.6) | 18.2 (0.7) | 25.4 (10.8) |
| Phone OS, N |  |  |  |  |  |  |  |
| Android | 0 | 7 | 12 | 40 | 35 | 69 | 163 (77%) |
| iOS | 16 | 4 | 0 | 19 | 4 | 5 | 48 (23%) |
| Gender, N |  |  |  |  |  |  |  |
| Female | 4 | 8 | 5 | 0 | 16 | 36 | 69 (33%) |
| Male | 12 | 3 | 7 | 57 | 23 | 38 | 140 (66%) |
| Missing | 0 | 0 | 0 | 2 | 0 | 0 | 2 (1%) |
| Education, N |  |  |  |  |  |  |  |
| High school | 16 | 1 | 12 | 0 | 39 | 74 | 142 (67%) |
| Associates | 0 | 6 | 0 | 3 | 0 | 0 | 9 (4%) |
| Bachelors | 0 | 3 | 0 | 36 | 0 | 0 | 39 (18%) |
| Graduate degree | 0 | 1 | 0 | 13 | 0 | 0 | 14 (7%) |
| Missing | 0 | 0 | 0 | 7 | 0 | 0 | 7 (3%) |
| Race/ethnicity, N |  |  |  |  |  |  |  |
| Non-Hispanic White | 7 | 9 | 9 | 32 | 14 | 46 | 117 (55%) |
| Non-Hispanic Black | 4 | 1 | 2 | 12 | 3 | 9 | 31 (15%) |
| Asian | 5 | 1 | 1 | 7 | 14 | 9 | 37 (18%) |
| American Indian | 0 | 0 | 0 | 0 | 0 | 2 | 2 (1%) |
| Other/Hispanic | 0 | 0 | 0 | 5 | 5 | 5 | 15 (7%) |
| Missing | 0 | 0 | 0 | 3 | 3 | 3 | 9 (4%) |

**Figure 1.**
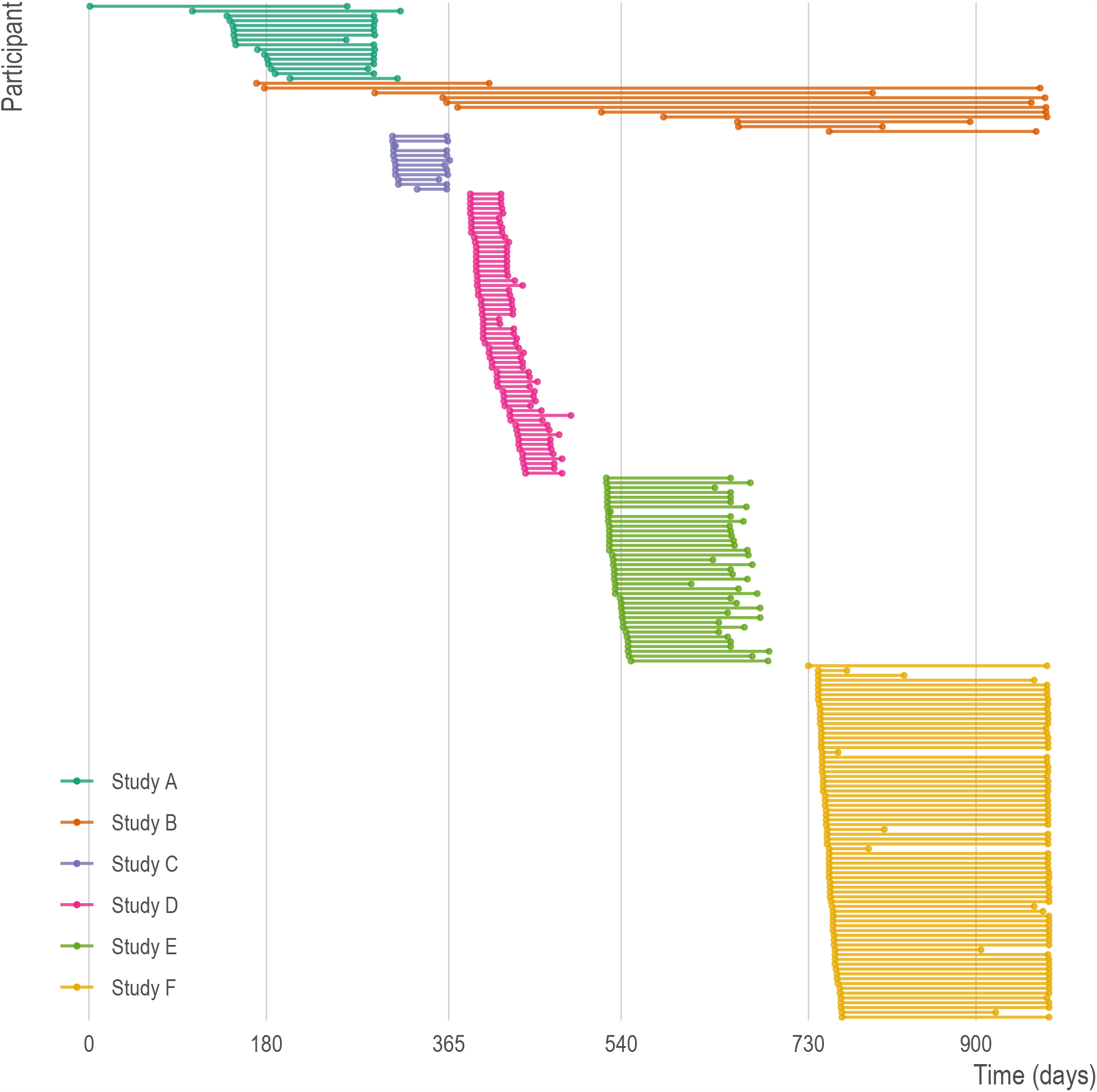
Periods of data collection for each study and each participant. Each horizontal line represents a single study participant with the endpoints at the first and last day of observation. Studies varied in number of participants, length of observation, and rate of attrition. Each study is represented by a different color. Note that because dates of study participation may be considered personally identifiable information, time (x-axis) is represented as days relative to the earliest date and not calendar time. All studies occurred between 2015 and 2018.

**Figure 2.**
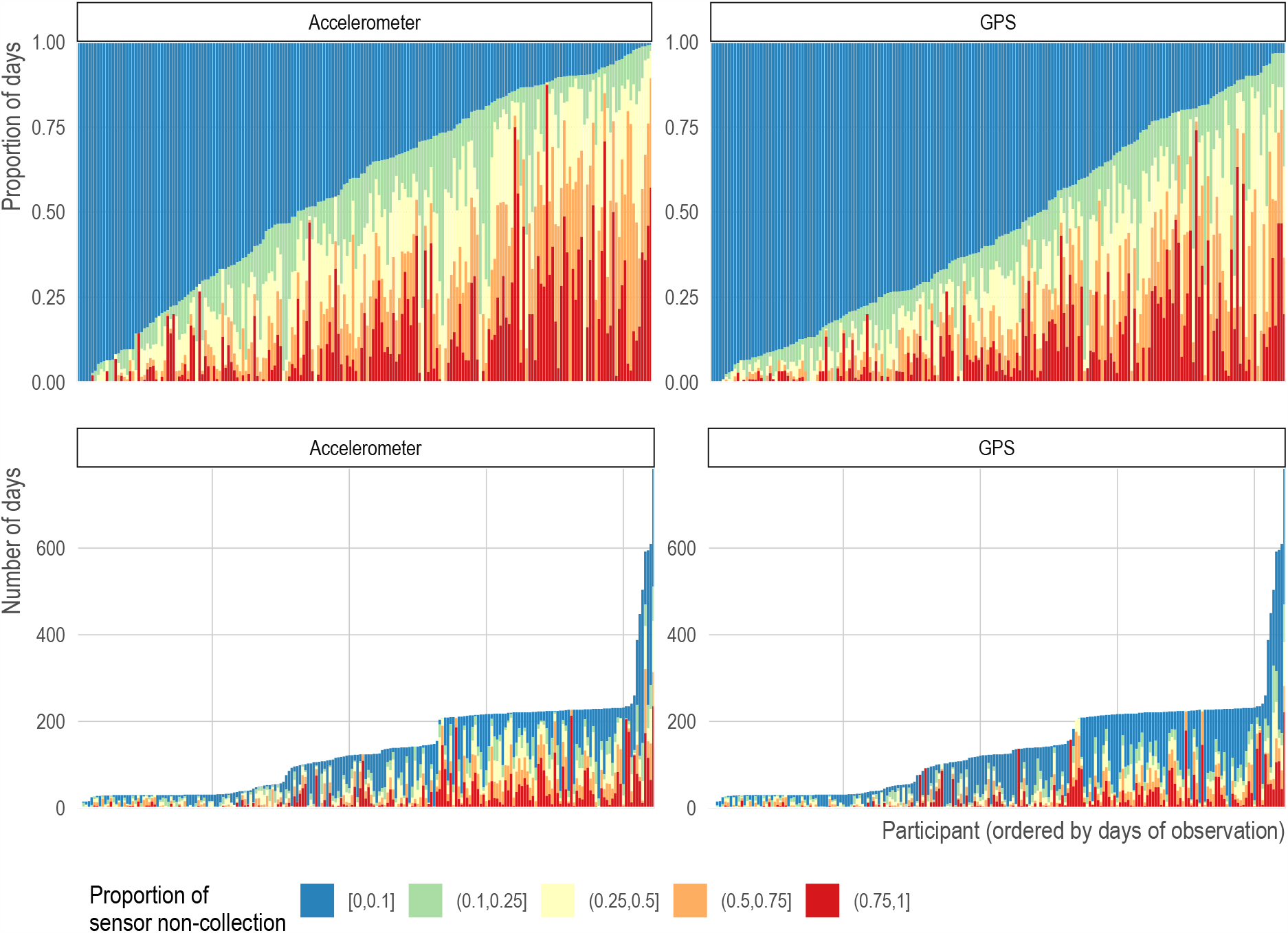
Days of observation by participant. Each vertical bar represents a single participant. In the top row, the height of the bar is the proportion of days of observations while the color reflects the proportion of daily sensor non-collection with blue representing low sensor non-collection and red representing high proportion of sensor non-collection. In the bottom row, the height of the bar is the absolute number of days. Participants are ordered by total number of days of observation. Follow-up was pre-specific in each study protocol based on time (i.e., not by the amount of data collected per subject).

Five of the six studies were conducted in the state of Massachusetts with four studies comprised of undergraduate students at Harvard College (Studies A, D, F, and G); one study involved patients known to be at risk for mania and psychosis from McLean Hospital (Study B); and one study (Studies E) consisted of an all-female subset of medical professionals in the Nurses’ Health Study 3^32^ with no diagnosed medical conditions. Study E is based in Massachusetts, but participants resided in several U.S. states. Each study received institutional review board (IRB) approval from their respective institutions for data collection (Table S2); another IRB approved by Harvard University governed the secondary analysis of the collected Beiwe data. Common inclusion criteria across all studies were: (1) ability to understand the English written consent form, (2) provision of written informed consent, (3) age 18 years or older, (4) possession of an Android or iOS smartphone, and (5) willingness to install the Beiwe application on their primary personal phone. Additional study-specific inclusion/exclusion criterion are listed in Table S2.

We investigated the role of various sociodemographic characteristics for rates of sensor non-collection using Bayesian hierarchical negative binomial models detailed in Methods. These models account for the correlated and nested nature of the data (i.e., observations within participants) and, unlike Poisson regression, allow for overdispersion of the data. The conditional average rates of sensor non-collection at the beginning of the studies were 14.2% (95% credible interval [CI]: 9.4, 21.3) for accelerometer and 25.0% (95% CI: 17.4, 35.6) for GPS (Table 2).

**Table 2.**
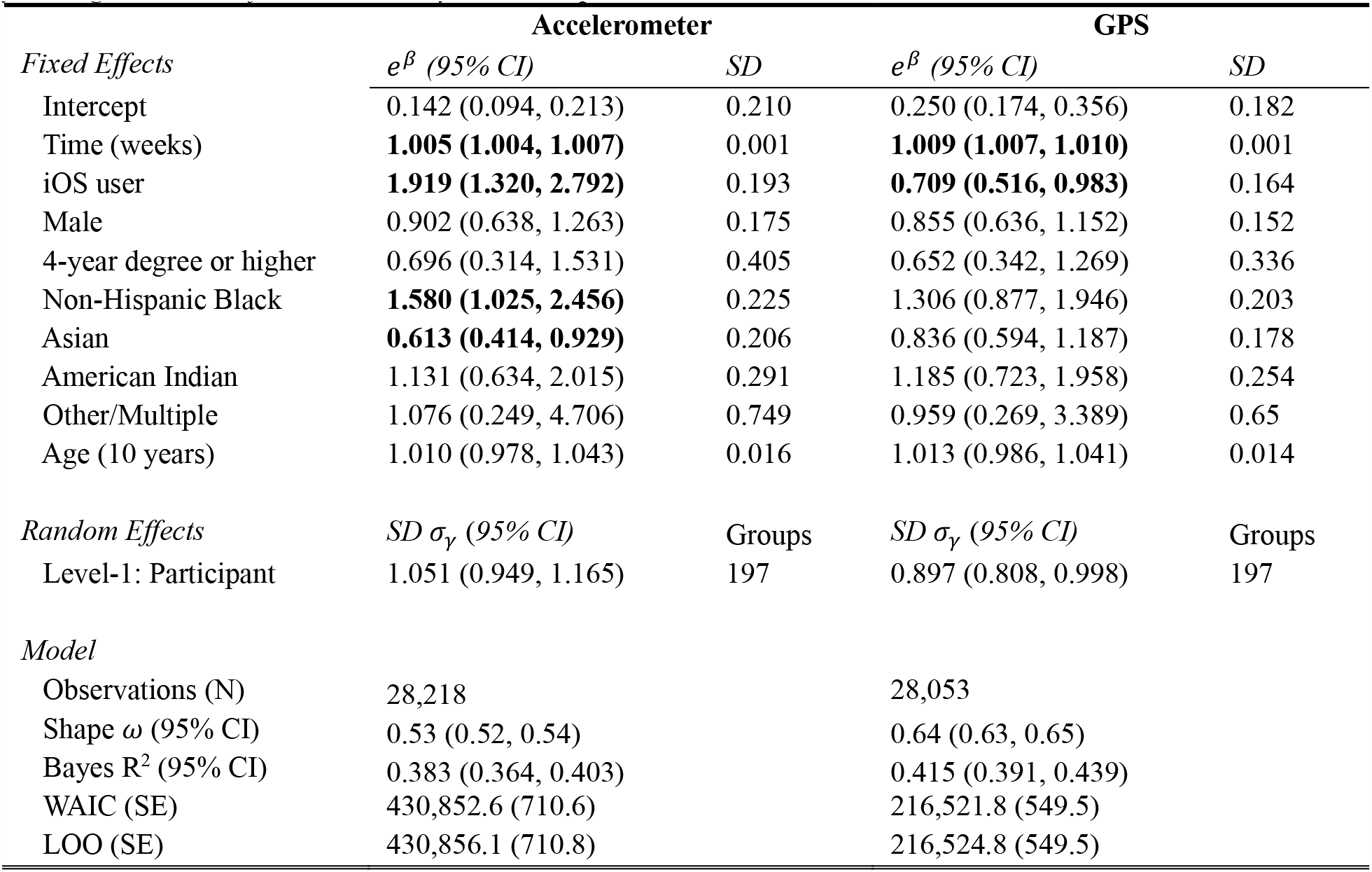
Model results. Model estimates for all parameters for sensor non-collection rates of accelerometer (left) and GPS (right). The coefficients and 95% credible intervals (95% CI) have been exponentiated to assist interpretation. Parameters with 95% CIs that exclude 1 are in bold. The reference group for education is less than 4-year degree and that for race/ethnicity is non-Hispanic White.

The rates of sensor non-collection increased over time at approximately 0.5% (95% CI: 0.4, 0.7) per week for accelerometer and 0.9% (95% CI: 0.7, 1.0) per week for GPS (Table 2). Participants with iOS devices had substantially higher rates of accelerometer non-collection (adjusted relative rate [RR]: 1.92 [95% CI: 1.31, 2.79]) and lower rates of GPS non-collection (RR: 0.71 [95% CI: 0.52, 0.98]) compared to participants with Android devices (Figure 3).

**Figure 3.**
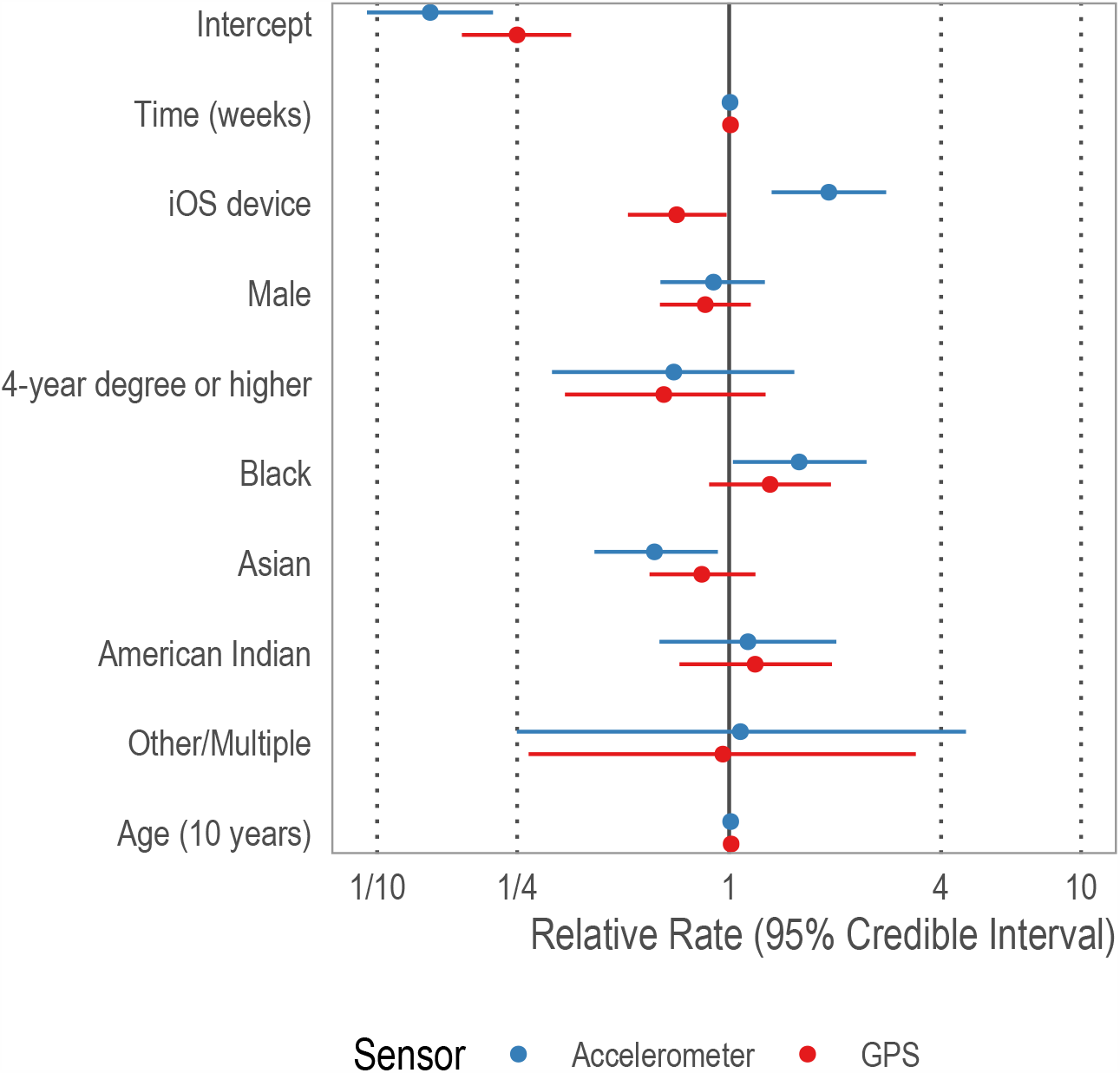
A forest plot of fixed effect estimates. The fixed effect estimates for accelerometer are in red and GPS in blue. Estimates have been exponentiated and can be interpreted as the relative change in sensor non-collection. The reference group for education is less than 4-year college degree and for race/ethnicity is non-Hispanic White. In terms of demographic characteristics, Black participants had higher rates of accelerometer non-collection compared to White participants; Asian participants had lower rates of accelerometer non-collection compared to White participants. iOS users had lower rates of GPS non-collection but higher rates of accelerometer non- collectin, suggesting systematic differences in the phone operating systems of each phone.

In terms of accelerometer non-collection and demographic characteristics, there was no significant difference between male and female participants (RR: 0.90 [95% CI: 0.64, 1.26]) or participants with a four-year college degree compared to those without (RR: 0.696 [95% CI: 0.31, 1.53]). Similarly, rates of accelerometer non-collection did not increase with age (RR: 1.01 [95% CI: 0.98, 1.04]). Compared to White participants, Black participants had approximately 58% (95% CI: 2, 146) higher rates of accelerometer non-collection, albeit with substantial uncertainty. Asian participants had lower rates of accelerometer non-collection (RR: 0.613 [95% CI: 0.414, 0.929]), again with substantial uncertainty. There was no similar difference for American Indian or Alaska Native participants (RR: 1.13 [95% CI: 0.63, 2.02]), or participants of other racial/ethnic descent (RR: 1.08 [95% CI: 0.25, 4.71]). Unlike accelerometer, there were no statistically significant racial/ethnic differences in rates of GPS non-collection. With the exception of phone type mention above, there were no differences across any of the demographic characteristics for GPS non-collection: gender, race/ethnicity, education, or age (Table 2 and Figure 3).

Compared to other model specifications, the selected models provide the best goodness-of-fit while remaining parsimonious (Supplementary Information Text S2). Using Bayes R^2^, the proposed models explain 38% (95% CI: 36, 40) of the variance in the rate of accelerometer non-collection and 42% (95% CI: 39, 44) of the variance in the rate of GPS non-collection (Table 2). Additionally, individual-level variation was substantial for both accelerometer (*σ*_*γ*_: 1.05 [95% CI: 0.95, 1.17]) and GPS (*σ*_*γ*_: 0.897 [95% CI: 0.81, 0.998]) non-collection (Figure 4). This level of between-individual variation implies the 25^th^ percentile participant would have about one-third lower accelerometer non-collection compared to the 75^th^ percentile participant (8% vs 26%) and one-quarter lower GPS non-collection (14% vs 53%).

**Figure 4.**
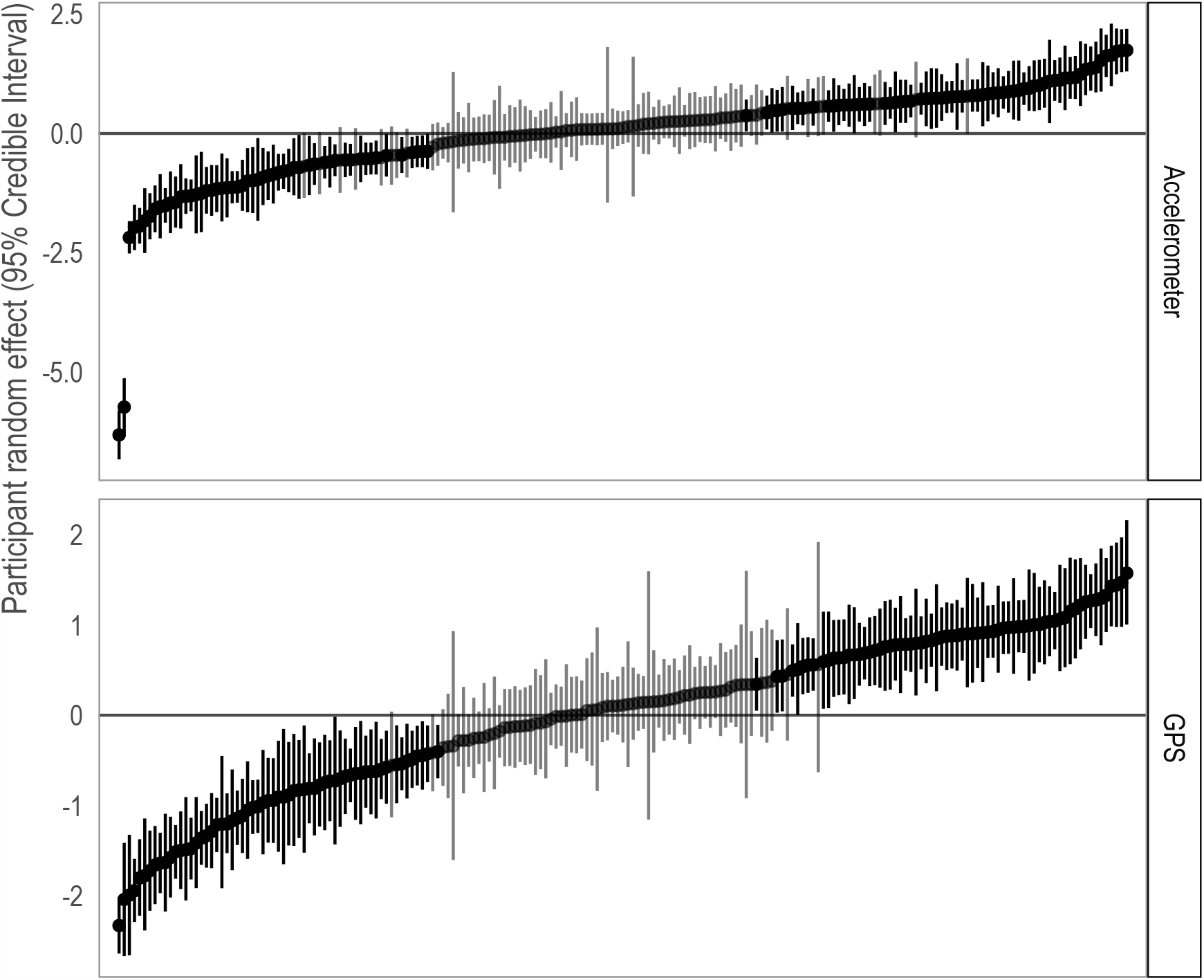
Participant random effect estimates for accelerometer (top) and GPS (bottom) by participant. The dots are the mean random effect estimates and the bars are the 95% credible intervals for each participant. Credible intervals that include 0 are shaded in grey while those that exclude 0 are shaded in black. The values on the y-axis represent the deviation from the overall average rate of sensor non-collection. There is substantial participant-level variation in missingness. In both panels, data have been ordered from lowest (i.e., least sensor non-collection) to highest (most sensor non-collection) median value. This level of between-individual variation implies the 25^th^ percentile participant would have about one-third lower accelerometer non-collection compared to the 75^th^ percentile participant (8% vs 26%) and one-quarter lower GPS non-collection (14% vs 53%). Note that the y-axes differ across the panels.

## Discussion

Our results suggest that overall sensor non-collection rates are 14% for accelerometer non- and 25% for GPS, with higher accelerometer non-collection and lower GPS non-collection among iOS users. In general, sensor non-collection did not vary by gender, age, or education.

Accelerometer non-collection among Black participants is slightly higher and accelerometer non-collection among Asian participants is slightly lower relative to White participants, and no racial/ethnic differences were observed for GPS non-collection. Importantly, while there is a statistically significant temporal trend of increasing sensor non-collection, the size of the effect is small (∼0.5–0.9% per week) and unlikely to be consequential in most studies relative to the baseline level of sensor non-collection. Lastly, we find large variation in the amount of sensor non-collection at the participant-level.

Our results have important implications for the design and analysis of future digital phenotyping studies. First, we show there is a nontrivial level of sensor non-collection across a variety of study settings and demographic characteristics. Future work in digital phenotyping needs to account for sensor non-collection through the development of new statistical methods, a better understanding of the reasons for sensor non-collection at the individual-level, and more reliance on within-subject over time study designs and data analyses that leverage the high adherence and long data collection periods of digital phenotyping. Similarly, researchers should account for the level of sensor non-collection when performing power calculations and recruiting participants by either recruiting a greater number of participants to offset potential missing data or by leveraging within-subject designs and planning for a longer period of follow-up. Some research questions may necessitate high-density, continuous GPS or accelerometer data, in which case it is often more statistically efficient to utilize a within-subject design with longer follow-up than a wide range of participants with limited follow-up.^37^

Second, we found substantial individual-level variability in sensor non-collection. This finding suggests that the observed large differences in sensor non-collection are not due to systematic study-related issues but are rather due to high between-person variability. The missingness appears to be independent of our measured, common demographic characteristics, and despite known differences in smartphone usage, it appears these differences in usage do not result in differential data collection in our sample. We note that, as with any study, there may be unobserved individual characteristics associated with missingness and thus detailed measurement of individual demographic factors is necessary to evaluate how missingness may affect specific outcomes of interest. Unmeasured, but likely important, individual-level factors include age or lifetime use of the phone and battery, charging habits, leisure activities such as hiking, camping, or other activities with where phone use is diminished. Such factors warrant future research.

Our study has several limitations. First, despite a large number of raw data measurements, measurement groupings, and person-days of observation, our sample still consisted of only six studies and 211 participants. We estimated few statistically significant associations between missingness and demographic characteristics, but this finding could potentially be explained by lack of statistical power. This is the largest meta-study of digital phenotyping data collection; however, as digital phenotyping studies move beyond the pilot stage, similar meta-study approaches to understanding missingness across important sociodemographic covariates will continue be necessary. Similarly, the heterogeneity of participants across studies and homogeneity within studies may drive some of our findings. For example, 12 of the 33 black participants come from a single study of female nurses. Thus, it is possible that our observed increased missingness among black participants is driven, at least in part, by occupation-related phone behaviors rather than by race/ethnicity. Differences between Android and iOS may be due to differences in the underlying userbase rather than software differences. In particular it appears that there may be a large socioeconomic difference between users of iOS and Android devices.

Non-scientific market surveys have consistently found higher self-reported income among iOS users compared to Android users,^38^ with one recent study reporting annual average salaries of approximately $53,000 and $37,000 for these two groups, respectively.^39^ Previous market research suggests Black Americans are more likely to own Android devices than their White counterparts.^40^ Fisher’s exact tests found no statistically significant differences between Android and iOS users across race/ethnicity or education in our data. Some subgroups may be more likely to own the latest phone and therefore own phones with greater battery capacity. Despite these limitations, we believe our study is informative for future digital phenotyping studies. In summary, we believe our results indicate digital phenotyping is feasible across a large and diverse sample when coupled with careful study design and statistical analysis.

## Methods

### Defining measurement groupings

The Beiwe Research Platform allows researchers to specify a sampling schedule separately for each sensor by adjusting the duration of the corresponding on-cycle and off-cycle. Using this information, we calculate the expected number of times the application attempts to collect data and the expected duration of data collection each day. However, ultimately the phone operating system controls data collection during an on-cycle, and considers factors such as battery life and computational load when making this determination. To account for these design considerations, we aggregated the raw measurements into “measurement groupings,” which we defined as chunks of data that were collected within a researcher-specified on-cycle and were separated from the next measurement grouping by at least half of the researcher-specified off-cycle (Table S2; Figure 2). Conceptually, a measurement grouping is an attempt by the smartphone application to collect data over some time interval, and it may have no observations (e.g., GPS was disabled by the participant) to several thousand (e.g., accelerometer data collection during a period of physical activity, such as running). Therefore, a missing measurement grouping (i.e., one with no observations), or sensor non-collection, could be due to (1) power management (e.g., low battery, a higher priority application is running, or high computational load); (2) sensor was disabled (e.g., activating airplane mode or deactivating GPS); or (3) the phone is off.

### Analysis

We used Bayesian hierarchical negative binomial regression to estimate the rate of sensor non-collection for GPS and accelerometer data. Unlike Poisson regression, negative binomial models allow for modeling both the mean and variance separately (i.e., allowing overdispersion), while the hierarchical framework accounts for the nested structure of the data (i.e., observations are clustered within users over time). For each user *i* in study *j*, the distribution of the rate of sensor non-collection per day *y*_*ij*_ is assumed to follow a negative binomial distribution. The mean of this distribution *μ*_*ij*_ is estimated as a log-linear function of *p* individual-level covariates *X*_1*ij*_ … *X*_*pij*_ with an study-specific offset *E*_*j*_, the expected number of measurement groupings per day (a known, fixed value that results from the specification of on-cycle and off-cycle for each sensor). Further, due to the non-independence of daily observations within each user, we allow for a user-specific random intercept *γ*_0*ij*_. The model can be written as

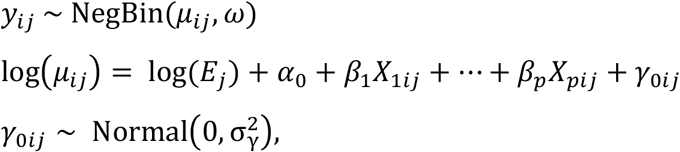

where the negative binomial distribution is parametrized in terms of the mean *μ*_*ij*_ and inverse overdispersion parameter ω.^41^ Here *α*_0_ is the grand mean across all individuals and *γ*_0*ij*_ is the individual-specific deviation from the grand mean. This individual-level random effect is assumed to be normally distributed with a mean of zero. The variance parameter of the random effect, 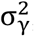, summarizes the variation in the rate of sensor non-collection at the individual level, after accounting for covariates. In addition, we estimated the fixed effects *β*_*p*_ using covariates *X*_*pij*_ at the individual level: duration in the study (in days), an indicator variable for operating system (iOS vs. Android), self-identified gender (male or female), educational attainment (less than four-year college degree or four-year degree and higher), race/ethnicity (non-Hispanic White, non-Hispanic Black, Asian, other race/multiple race/Hispanic, or American Indian / Alaskan Native), and age. We assume a common variance across studies; however, sensitivity analyses presented in Supplementary Information Text S2 indicate our results are robust to several model specifications.

Models were fit using the No-U-Turn Sampler,^42^ an efficient, adaptive Hamiltonian Monte Carlo algorithm. Specifically, we used the `brm()` function from the `brms` package^43^ which interfaces with the Stan library.^44^ All parameters were assigned the default brms priors.

Specifically, fixed effects were assigned an uninformative, improper prior *β*∼Uniform(−∞, +∞); the intercept was assigned the diffuse prior *α*∼Student^’^s *t*(3, 6.7, 2.5); and the standard deviation of the random effects were assigned the diffuse prior *σ*_*γ*_ ∼Half − Student^’^s *t*(3, 0, 2.5). All models were fit using eight independent chains. Model convergence was assessed using the rank-normalized-split-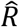 and rank-normalized-folded-split-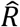, and the model was considered successfully converged when the maximum of both 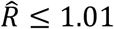. To ensure reliable posterior estimates, each chain was run until the Bulk Effective Sample Size and Tail Effective Sample Size metrics reached at least 100 samples per chain (Supplementary Information).^45^ We used the widely applicable information criterion (WAIC),^46^ the asymptotically-equivalent leave-one-out cross-validation^47^ with Pareto smoothed importance sampling (LOO),^48^ and Bayesian R-squared (Bayes *R*^2^)^49^ to evaluate model goodness-of-fit, the necessity of random effects components, other nesting structures (e.g., observations within users within studies or observations within studies), and other model specifications (Supplementary Information Text S2). All analyses were performed in R 4.0.2.^50^

### Availability of data, materials, and methods

While this research does not used only metadata (e.g., timestamps of GPS pings rather than coordinates), dates of participant activity can be considered personally identifiable information; therefore, the data cannot be shared publicly. Data available upon request, contingent upon appropriate IRB approvals or exemptions from participating institutions. While not the raw data, these data will provide sufficient information to reproduce our results (e.g., using shifted and/or adding noise to timestamps, re-randomized user identifiers). Replication code can be found at https://github.com/mkiang/beiwe_missing_data or https://github.com/onnela-lab/beiwe_missing_data (Supplementary Information Text S3). The Beiwe platform is open source and publicly available (Supplementary Information Text S1).

## Acknowledgements

We would like to thank Timothy O’Keefe for providing technical expertise and assistance. In addition, Jeanette Lorme and Maria Simoneau provided project support.

## Author Contributions

JPO and MVK designed the study. JPO, JTB, RLB, GC III, and JWR-E acquired the data. MVK did the data analysis. JTC and MJA suggested additional analyses as appropriate. All authors interpreted the results. MVK, JPO, and JTC drafted the manuscript. All authors provided critical revisions to the manuscript. All authors reviewed the manuscript and approved the final version to be published.

