## Supplemental Materials for "Sociodemographic Characteristics of Missing Data in Digital Phenotyping"

### Supplementary Information for *Sociodemographic Characteristics of Missing Data in Digital Phenotyping*

### Supplementary Tables

#### Table S1. Summary statistics of sensor data by study.

*Data are from six studies with a wide range in number of participants, time in study (person-days), number of raw data points collected (Obs.), number of measurement groupings (M.G.s), and percent of sensor non-collection (N.C.). Note that both Obs. and M.G.s are expressed in millions (M). For Studies A, B, C, E and F, the accelerometer on/off cycles were set for 10 seconds on / 10 seconds off. For Study D, the accelerometer on/off cycle was set for 10 seconds on / 1200 seconds off. For GPS, the on/off cycles, in seconds, were: Study A and C (90/1200); Study B and F (120/600); Study D (60/1200); and Study E (60/300).*

| | **Accelerometer** | | | | **GPS** | | | |
|---|---|---|---|---|---|---|---|---|
| | *Person-days (#)* | *Obs.* | *M.G.* | *N.C. (%)* | *Person-days (#)* | *Obs.* | *M.G.* | *N.C. (%)* |
| Study A | 2200.56 | 979.89M | 8.87M | 24.11 | 2122.81 | 10.93M | 0.17M | 33.73 |
| Study B | 4753.72 | 1186.09M | 12.91M | 38.24 | 4760.00 | 37.75M | 0.44M | 33.36 |
| Study C | 551.89 | 177.03M | 1.42M | 41.54 | 551.94 | 4.60M | 0.10M | 28.35 |
| Study D | 1893.02 | 13.96M | 0.10M | 28.78 | 1878.74 | 5.10M | 0.11M | 27.60 |
| Study E | 4667.43 | 1583.03M | 14.13M | 30.76 | 4643.05 | 14.89M | 0.32M | 22.54 |
| Study F | 15442.52 | 4206.92M | 41.55M | 38.11 | 15390.42 | 39.51M | 0.97M | 28.27 |
| **Total** | **29509.14** | **8146.92M** | **78.98M** | **35.37** | **29346.95** | **112.77M** | **2.12M** | **28.53** |

#### Table S2. Institutional review board approval and inclusion/exclusion criteria.

*Institutional review board (IRB) approval was granted for data collection for each study, by their respective institutions, with the following inclusion/exclusion criteria. In addition, for all studies, IRB approval was granted for the secondary data analysis to be performed at Harvard TH Chan School of Public Health (IRB16-0966).*

| **Study** | **IRB Protocol Number (Institution)** | **Additional Inclusion/Exclusion Criteria** |
|---|---|---|
| A and F | IRB16-1230 (Harvard) | Inclusion: Normal, cognitively intact college students; 18-28 years old; native of fluent English speakers; recruitment focused on freshmen |
| C | 2015P002189 (Massachusetts General Hospital) | Inclusion: Must have severe affective and psychotic illness; over the age of 18 |
| D | IRB16-0440 (Harvard) | Inclusion: Normal, cognitively intact college students; 18-22 years old; native of fluent English speakers; students enrolled in exam-based science courses (any grade)<br>Exclusion: Contraindications for MRI |
| E | 2006P000473 (Brigham and Women's Hospital) | Inclusion: Must be a current participant of the Nurses' Health Study 3. |
| G | IRB15-3613 (Harvard) | Inclusion: Normal, cognitively intact college students; 18-22 years old; native of fluent English speakers; students enrolled in exam-based science or math courses (any grade) |

### Supplementary Figures

#### Figure S1. Periods of data collection for each study and each participant.

*Each horizontal line represents a single study participant with the endpoints at the first and last day of observation and the intensity of color represents the amount of data collection per day such that darker tones have higher sensor non-collection (i.e., more missing data). Studies varied in number of participants, length of observation, and rate of attrition. Each study is represented by a different color. Note that because dates of study participation may be considered personally identifiable information, time (x-axis) is represented as days relative to the earliest date and not calendar time. All studies occurred between 2015 and 2018.*

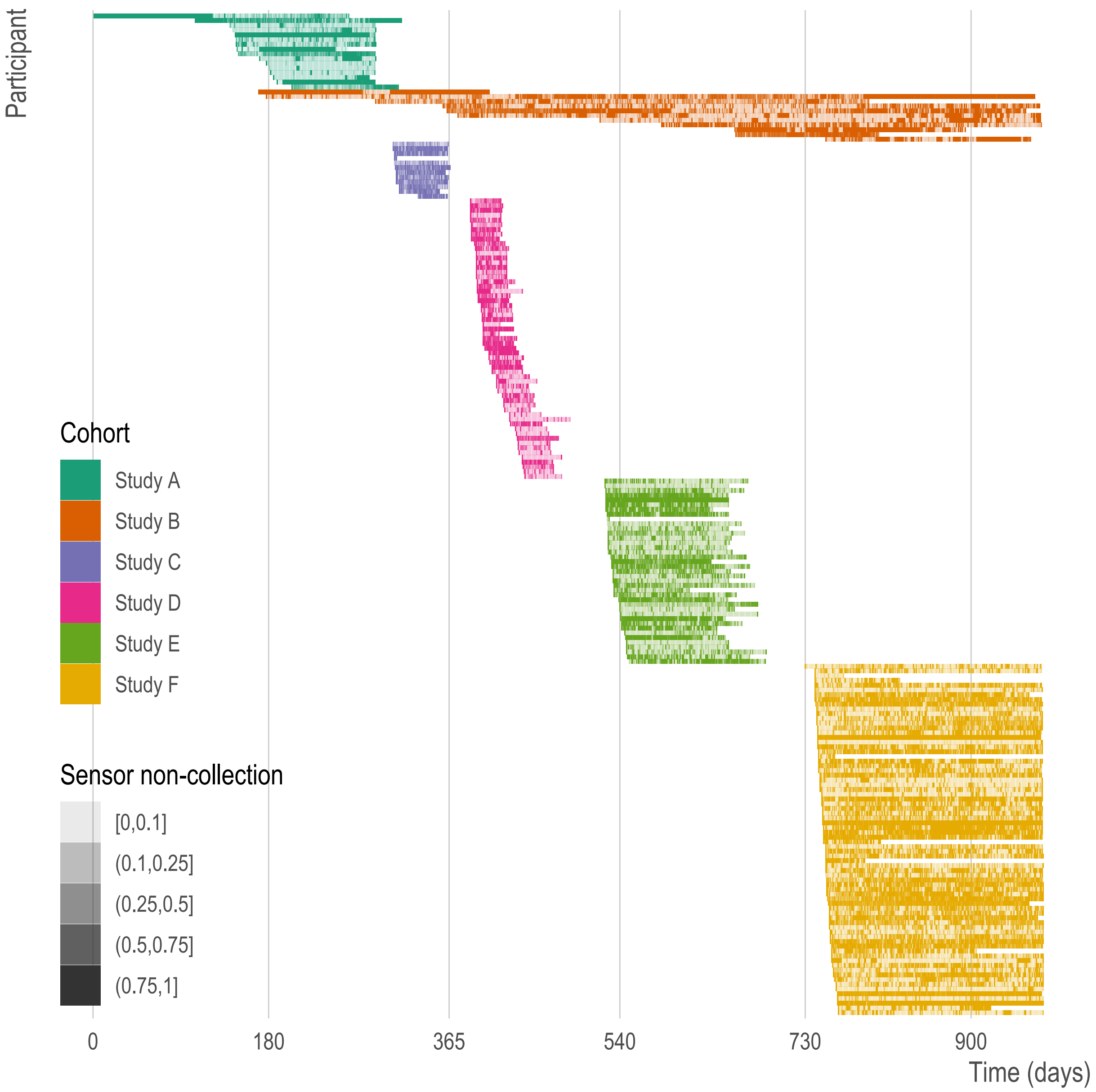

### Supplementary Text

##### Text S1. Open-source code repositories for Beiwe.

There are three affiliated code repositories for the Beiwe Research Platform. The Beiwe Research Platform is powered by backend code that can be found at `https://github.com/onnela-lab/beiwe-backend`. The Beiwe Android frontend code can be found at `https://github.com/onnela-lab/beiwe-android`. The Beiwe iOS frontend code can be found at `https://github.com/onnela-lab/beiwe-ios`. More information about Beiwe can be found at `https://www.beiwe.org`.

##### Text S2. Assessing goodness-of-fit with alternative model specifications.

Given the structure of the data, several alternative models (with different assumptions) can be reasonably specified: (1) a non-hierarchical model, (2) a non-hierarchical model with study fixed effects, (3) a three-level hierarchical model with observations nested in users nested in studies, (4) a two-level hierarchical model with observations nested in users and study fixed effects, and (5) a two-level hierarchical model with observations nested in study. Here, we compare these alternative model specifications to the model presented in the paper (two-level model with observations nested in users).

Consistent with the model in the main text, assume for each user $i$ in study $j$, the rate of sensor non-collection per day $y_{ij}$ follows a negative binomial distribution. The mean rate of sensor non-collection $\mu_{ij}$ is estimated as a log-linear function of $p$ covariates $X_{1ij} \dots X_{pij}$ with a study-specific offset $E_j$, the expected number of measurement groupings per day (a known, fixed, value). Further, due to the non-independence of daily observations within each user, we allow for a user-specific random intercept $\gamma_{0ij}$. Here, we additionally add a study-specific random intercept $\delta_{0j}$. This model can be written as

$$y_{ij} \sim \text{NegBin}(\mu_{ij}, \omega)$$

$$\log(\mu_{ij}) = \log(E_j) + \alpha_0 + \beta_1 X_{1ij} + \cdots + \beta_p X_{pij} + \gamma_{0ij} + \delta_{0j}$$

$$\gamma_{0ij} \sim \text{Normal}(0, \sigma_\gamma^2)$$

$$\delta_{0j} \sim \text{Normal}(0, \sigma_\delta^2),$$

where the negative binomial distribution is parametrized in terms of the mean $\mu_{ij}$ and inverse overdispersion parameter ω. Note that the primary model presented in the manuscript is equivalent to the above specification when setting $\sigma_\delta^2 = 0$. A reasonable, simpler specification would be to set both $\sigma_\delta^2$ and $\sigma_\gamma^2$ equal to zero for a non-hierarchical model. This non-hierarchical specification could also include study fixed effects in addition to the covariates outlined in the manuscript. Similarly, we could specify the same model as shown in the paper but add study fixed effects (i.e., as covariates). Alternatively, we could specify a two-level model (as done in the manuscript) but set $\sigma_\gamma^2 = 0$ so observations are nested in study (but not users). Again, we could do this with and without study fixed effects. Lastly, we could nest observations in users and users in study, allowing all the above parameters to be estimated. A summary of these models is shown below.

| | User random effect | Study random effects | Study fixed effects | Accelerometer | | GPS | |
|---|---|---|---|---|---|---|---|
| | | | | Δ LOO (SE Δ) | Δ WAIC (SE Δ) | Δ LOO (SE Δ) | Δ WAIC (SE Δ) |
| Non-hierarchical model | | | | -3,476.4 (90.7) | -3,478.4 (90.4) | -3,828.3 (95.3) | -3,829.7 (95.2) |
| Non-hierarchical model with study fixed effects | | | X | -3,280.4 (88.4) | -3,281.9 (88.1) | -3,725.7 (94.5) | -3,738.0 (94.0) |
| **Two-level model with user random effects** | **X** | | | **-0.3 (0.7)** | **-0.0 (0.0)** | **-0.2 (0.5)** | **-0.2 (0.5)** |
| Two-level model with user random effects and study fixed effects | X | | X | 0.0 (0.0) | -0.7 (0.8) | 0.0 (0.0) | 0.0 (0.0) |
| Two-level model with study random effects | | X | | -3,280.5 (88.4) | -3,282.0 (88.1) | -3,736.7 (94.5) | -3,737.8 (94.4) |
| Three-level model with user and study random effects | X | X | | -0.3 (0.4) | -0.4 (0.6) | -0.5 (0.3) | -0.5 (0.3) |

For each model, we calculated the goodness-of-fit using Pareto smoothed leave-one-out cross-validation (LOO) and the asymptotically-equivalent widely applicable information criterion (WAIC). For each sensor, we calculated the difference in the theoretical expected log pointwise predictive density of each model relative to the best performing model (ΔLOO and

ΔWAIC) and the standard error of this quantity (SE Δ). As shown above, the three-level model with user and study random effects, the two-level model with user random effects and study fixed effects, and the primary model (two-level with user random effects) performed substantially better than the alternative parameterizations. For both sensors, the primary model used in the manuscript was either the best performing model or was within 1 standard error of the best performing model. We selected this model because it is the most parsimonious model; however, we note that parameter estimates do not change when using the other two specifications. See the online supplement for all model estimates (see also Text S3).

#### Text S3. Data availability and replication code.

While the data used in these analyses were *meta*data and contained only timestamps (e.g., the date and time of a GPS ping but not the coordinates), the timestamps of participants can be considered personally identifiable information. Therefore, to minimize the potential for participant harm and re-identification, the data are not shared publicly. We will, however, make data available upon request with sufficient information to reproduce our results (e.g., using shifted and/or adding noise to timestamps). In addition, we provide example replication code, along with documentation, in an online repository. The code and documentation are near exact copies of the code used in this project with only minor differences. Specifically, for this paper, we use internal study project names which may include a year and/or month. Out of an abundance of caution, we remove any references to these study names. However, the code is otherwise the same. Please see the repository at: `https://github.com/mkiang/beiwe_missing_data` or `https://github.com/onnela-lab/beiwe_missing_data`.
